# Health Catch-UP!: a realist evaluation of an innovative multi-disease screening and vaccination tool in UK primary care for at-risk migrant patients

**DOI:** 10.1101/2024.06.13.24308888

**Authors:** Jessica Carter, Lucy P Goldsmith, Felicity Knights, Anna Deal, Subash Jayakumar, Alison F Crawshaw, Farah Seedat, Nathaniel Aspray, Dominik Zenner, Philippa Harris, Yusuf Ciftci, Fatima Wurie, Azeem Majeed, Tess Harris, Philippa Matthews, Rebecca Hall, Ana Requena-Mendez, Sally Hargreaves

**Affiliations:** Migrant Health Research Group, Institute for Infection and Immunity, St George’s, University of London; Population Health Research Institute, St George’s, University of London; Faculty of Public Health and Policy, LSHTM; The Stonebridge Practice, Harness PCN South, NHS North West London Integrated Care System; Wolfson Institute of Population Health, Queen Marys University of London; Department of Primary Care and Public Health, Imperial College London; Addiction and Inclusion Directorate, Office for Health Improvement and Disparities, Department of Health and Social Care, 39 Victoria Street, London, SW1H 0EU; Barcelona Institute for Global Health (IS Global Campus Clinic), Spain; Islington GP Federation, London; Guy’s and St Thomas’ NHS Foundation Trust; Clinical Research Department, London School of Hygiene and Tropical Medicine and Division of Infection, UCLH; Experts by Experience (advisor)

**Keywords:** Migrant health, infectious disease, non-communicable disease, screening, primary care, clinical decision support tool, digital solutions, multi-disease, vaccination

## Abstract

**Background:** Migrants to the UK face disproportionate risk of infections, non-communicable diseases, and under-immunisation compounded by healthcare access barriers. Current UK migrant screening strategies are unstandardised with poor implementation and low uptake. Health Catch-UP! is a collaboratively produced digital clinical decision support system that applies current guidelines (UKHSA and NICE) to provide primary care professionals with individualised multi-disease screening (7 infectious diseases/blood-borne viruses, 3 chronic parasitic infections, 3 non-communicable disease or risk factors) and catch-up vaccination prompts for migrant patients, which needs evaluating as a complex intervention to explore effectiveness and acceptability.

**Methods:** We carried out a mixed-methods process evaluation of Health Catch-UP! in two urban primary healthcare practices to integrate Health Catch-UP! into the electronic health record system of primary care, using the Medical Research Council framework for complex intervention evaluation. We collected quantitative data (demographics, patients screened, disease detection and catch-up vaccination rates) and qualitative participant interviews to explore acceptability and feasibility.

**Results:** 99 migrants were assessed by Health Catch-UP! across two sites (S1, S2). 96.0 % (n=97) had complete demographics coding with Asia 31.3 % (n= 31) and Africa 25.2% (n=25) the most common continents of birth (S1 n=92 [48.9% female (n=44); mean age 60.6 years (SD 14.26)]; and S2 n=7 [85.7% male (n=6); mean age 39.4 years (SD16.97)]. 61.6% (n=61) of participants were eligible for screening for at least one condition and uptake of screening was high 86.9% (n= 53). Twelve new conditions were identified (12.1% of study population) including hepatitis C (n=1), hypercholesteraemia (n= 6), pre-diabetes (n=4) and diabetes (n=1). Health Catch-UP! identified that 100% (n=99) of patients had no immunisations recorded; however, subsequent catch-up vaccination uptake was poor (2.0%, n=1). Qualitative data supported acceptability and feasibility of Health Catch-UP! from staff and patient perspectives, and recommended Health Catch-UP! integration into routine care (e.g. NHS health checks) but required an implementation package including staff and patient support materials, standardised care pathways (screening and catch-up vaccination, laboratory, and management), and financial incentivisation.

**Conclusions:** Clinical Decision Support Systems like Health Catch-UP! can improve disease detection and implementation of screening guidance for migrant patients but require robust testing, resourcing, and an effective implementation package to support both patients and staff.

## Introduction

Migration has risen at an unprecedented level in recent years, with the numbers of labour migrants seeking work opportunities, asylum seekers and refugees, and people displaced by conflict, natural disasters, and climate change at their highest levels since records began.(1) Migrants are a diverse group, but compared to host populations in high income receiving countries such as the United Kingdom, are disproportionately impacted by a range of infectious diseases that are more common in their countries of origin, with implications for health care provision and wider public health.(2) Hence, in 2018 the European Centre for Disease Prevention and Control (ECDC) published evidence-based guidance highlighting the need to screen at risk migrant groups for tuberculosis (TB), HIV, Hepatitis B and C, schistosomiasis and strongyloidiasis, establishing a screening criteria based on country of origin, as well as recommending catch-up vaccinations to offer to child and adult migrants.(3) Migrants from some groups have also been shown to be at increased risk of several non-communicable diseases. These include diabetes, which develops earlier than in the host population, haemoglobinopathies such as sickle cell anaemia common in Sub-Saharan Africa, and cardiovascular and cerebrovascular disease dependent on country of origin, country of destination, and duration of residence. (4)

Recent work on integrated multi-disease screening (screening for more than one condition at one time point) suggests it is an effective strategy for migrant groups with the potential for better uptake, feasibility and acceptability compared to single disease screening programmes which have to date been the focus. (5–7) However, despite the evidence and policy suggesting the need for holistic assessment of risk factors and multi-disease screening in migrants after arrival, most countries do not implement any systematic screening, and those that do have historically only screened for tuberculosis. (8, 9) Additionally, most current screening interventions exist in specialised clinics often based in secondary care which risks missing a large proportion of the migrant population accessible through primary care.(9) Current screening interventions often fail to include an individualised assessment of risk based on demographics or the threshold level of prevalence for infectious diseases in the country of origin (the basis of many screening guidelines, such as NICE, UKHSA, ECDC).(7, 9)

This variation and assessment of risk for each disease depending on individual differences (gender, age, country of origin, duration of residence, etc) creates a practical and clinical challenge for clinicians, particularly in primary care, due to the combination of time pressures, workload, knowledge gap due to lack of provision of migrant health training and clinical infectious disease experience. (6, 9, 10) Many clinicians are unaware of the primary care guidance on which risk assessments can be based, summarised in **Table 1**.(9) Additionally, key demographic details affecting risk, for example country of origin and date of entry to the UK, are not routinely coded into electronic patient records in UK primary care. This limits our ability to detect gaps in screening and vaccination coverage and address screening and catch-up vaccination needs for specific migrant groups.(9–11) In other clinical areas facing such risk variation, clinical decision support systems (CDSSs) have been adopted. These use a computerised algorithm to assess a range of patient characteristics and provide tailored recommendations to support clinical decision making.(12) The use of CDSSs remains relatively novel in providing effective migrant care, however initial piloting of this approach to clinical support in Spain suggests high levels of feasibility, acceptability, and an increase in screening and disease detection.(13) In the UK, a CDSS called Health Catch-UP! has been developed in collaboration with primary care teams, patients with lived experience of migration, academics, infectious disease experts, digital software specialists (EMIS), and UKHSA. In this study we take a realist approach using process evaluation methodology to evaluate this CDSS in two primary care practices in North London with high migrant populations.

**Table 1:** Summary of Guidance Regarding Risk Assessment for Migrant Patients.

| Disease | Guidance | Recommendation / Patients defined as 'At Higher Risk' |
| --- | --- | --- |
| Vaccination | Public Health England (now UKHSA). Vaccination of individuals with uncertain or incomplete immunisation status. Updated 1<sup>st</sup> September 2023.<br>https://www.gov.uk/government/publications/vaccination-of-individuals-with-uncertain-or-incomplete-immunisation-status | The UKHSA advises to assume that patients are unimmunised if they are unable to provide reliable written or verbal vaccination history, and to offer vaccination according to the host country's vaccination schedule. In the UK this is to readminister MMR (2 doses), td/IPV (3 doses), and then to consider vaccines including MenACWY and HPV up to age 25 years. |
| HIV | HIV testing: increasing uptake among people who may have undiagnosed HIV (Joint NICE and Public Health England 2016) https://www.nice.org.uk/guidance/ng60 | Patients under 65 years from a higher prevalence country (Much of Africa, Asia & Caribbean) |
| Latent TB | Tuberculosis. National Institute for Health and Care Excellence. Clinical knowledge summaries. (2019)<br>Latent TB infection testing and treatment programme for migrants: Presenting data between 1 April 2015 to 31 March 2020. Public Health England. | NICE recommends that new entrants aged under 65 from high incidence countries, are screened for latent TB using interferon-gamma release assay via a single blood test.<br>In England, UKHSA's latent TB testing and treatment programme exists in primary care for people aged 18-35, who have arrived from high incidence countries within the last five years. |
| Hepatitis B and C | Hepatitis B and C testing: people at risk of infection<br>Public health guideline [PH43] | NICE recommends that GPs and practice nurses should test people born or brought up in a country with an intermediate or high prevalence (2% or greater) of chronic hepatitis B. This includes all countries in Africa, Asia, the Caribbean, Central and South America, Eastern and Southern Europe, the Middle East and the Pacific islands. |
| Strongyloidiasis | European Centre for Disease Prevention and Control. Public health guidance on screening and vaccination for infectious diseases in newly arrived migrants within the EU/EEA. (2018) | ECDC guidance recommends serological screening for strongyloidiasis, irrespective of number of years since leaving endemic countries, particularly in individuals who are immunosuppressed. |
| Schistosomiasis | European Centre for Disease Prevention and Control. Public health guidance on screening and vaccination for infectious diseases in newly arrived migrants within the EU/EEA. (2018) | ECDC suggests offering serological screening to all migrants from countries of high endemicity in sub-Saharan Africa, and focal areas of transmission in Asia, South America and North Africa. |
| Chagas Disease | The Migrant Health Guide, Office for Health Improvement and Disparities [Accessed 21/03/24] | Consider the possibility of Chagas disease in migrants from endemic South and Central American countries. |
| Haemoglobinopathy screening (Sickle Cell / Thalassaemia) | The Migrant Health Guide, Office for Health Improvement and Disparities [Accessed 21/03/24] | Recommends being alert to the possibility haemoglobin disorders which are most prevalent in tropical regions; thalassaemia more common in Asia, the Mediterranean basin and the Middle East, whilst sickle cell predominates in Africa. |
| Diabetes | Type 2 Diabetes: Prevention in People at High Risk (NICE 2012) | Risk assess all people aged 25 to 39 of South Asian, Chinese, African-Caribbean, black African and other high-risk black and minority ethnic groups, except pregnant women, as well as all adults aged 40 or above. |
| Lipid check (for cardiovascular disease) | Cardiovascular disease: risk assessment and reduction, including lipid modification<br>NICE guideline [NG238] | Review estimates of CVD risk on an ongoing basis for people over 40 (The NHS Health Check which includes lipid testing is offered every five years. Note this is not a specific check for migrants). |

## Methods

### Evaluation Design and Rationale

Realist evaluation is a flexible theory-driven but active approach embedded in the reality of changing contexts influencing intervention implementation and how the actors involved in implementation respond to these changes.(14) It allows consideration of the mechanisms *by* which and the circumstances *in* which programmes work for specific stakeholders.(14) We adopted the Medical Research Council (MRC) framework for the design and evaluation of complex interventions (**Appendix 1**) to provide insights into the context-mechanism-outcome interactions of the Health Catch-UP! tool in two primary care settings.(15) (16) (17) Implementation of the Health Catch-UP! tool is inherently a complex intervention due to the number of components involved, the range of behaviours targeted, and the interaction between the intervention and the context in which it is implemented.(18)

In our evaluation we aimed to generate core insights on the process and challenges of implementation of Health Catch-Up! to inform iterative modification of both the intervention and our underlying programme theory (the set of assumptions underlying an intervention that explains why the planned activities should lead to the predefined goals and objectives).(15) We therefore sought to retest and refine our programme theory whilst assessing whether and how Health Catch-Up! implementation was successful and report this evaluation in accordance with RAMESES II reporting standards for realist evaluations.(14) Our evaluation was split into two phases: phase one focused on development of the intervention and initial programme theory. Phase two focused on iteratively refining and evaluating Health Catch-UP! through a pilot implementation process evaluation (with no control group) focusing on formative rather than outcome valuations according to realist principles.(19)

### Intervention Description

The intervention is the integration of the CDSS Health Catch-UP! into the electronic health record (EHR) system of primary care, to support implementation of UK migrant health guidelines for infectious disease and selected non-communicable disease screening and catch-up vaccination. (20) (21, 22) (23) (24–28) (27)The tool works in two stages, the first stage requires the primary health care professional (PHCP) to ask and code six key demographic variables to ascertain risk (age, sex, body mass index (BMI), country of origin, ethnicity, and date of entry to the UK (which must be 4 years or fewer for LBTI screening)). In stage two, the demographic coded responses are integrated with existing coded clinical information including results of previous screening to produce a single “pop-up” or prompt which summarises the guideline-recommended screening blood tests and vaccines individualised to that patient. The PHCP is not prompted to order a screening test if tests have previously been done and results recorded on the patient’s electronic health record. Through this two-step process Health Catch-UP! facilitates the first routine data collection on migrant health in UK primary care.

Health Catch-UP! has been collaboratively developed with a multi-disciplinary team and EMIS – digital health specialists who provide the most widely used electronic patient record systems and software in primary care). We repeatedly drew on the knowledge of our stakeholder groups to inform the selection of which diseases to screen for within Health Catch-UP!, outlined below and how to prompt clinicians to offer these, with screening focused on a core set of communicable and non-communicable conditions as per UK guidelines (see Table 1). It was felt to be important that conditions could be tested for using a simple blood test and have the potential to not require an in-person doctor appointment. It was agreed that Health Catch-UP! should prompt the PHCP to use the tool through a small visual prompt or pop-up. These visual prompts or reminders are commonly used for other conditions, for example suggesting when patients should be offered a cervical smear test, and therefore PHCPs would be accustomed to seeing and actioning them.

Health Catch-UP! applies the UK guidelines (UKHSA migrant health guide and NICE guidelines) for screening for seven infectious diseases including the blood-borne viruses: HIV, hepatitis B and C, latent tuberculosis (LTBI), and three chronic parasitic infections: strongyloidiasis, schistosomiasis and Chagas disease, as well as three non-communicable diseases or risk factors: diabetes (tested through glycated haemoglobin: Hba1c), high cholesterol (a risk factor for cardiovascular disease) and haemoglobinopathy (sickle cell disease, thalassaemia). Health Catch-UP! also prompts healthcare staff to ask questions about immunisation status and offer catch-up vaccination to align all patients with the UK schedule. According to guidance, catch-up vaccinations should be part of routine care and include measles, mumps, rubella (MMR), tetanus, diphtheria, polio (Td/IPV), HPV (aged 11-25 years) and meningococcal (MenACWY) (aged 10-25 years) vaccines (**Table 1**; **Appendix 2**).(20)

### Phase 1 Methodology: Generation of the Intervention and Initial Programme Theory

The role of a programme theory model is to describe how an intervention is expected to lead to its effects and under what conditions this will happen. The team collaboratively developed an Initial Programme Theory (IPT) to form the basis of the evaluation and inform subsequent study design, data collection and analysis. This was refined iteratively as our understanding progressed. We then interviewed 64 UK-based clinical and non-clinical primary care professionals to explore their views on the context and function of current infectious disease screening and adult catch-up vaccination processes, and to the intervention Health Catch-UP! including barriers and facilitators to implementation. We modified and refined our initial theory based on these data (published elsewhere in full).(9) (10)

### Phase 2 Methodology: Pilot Implementation and Evaluation

#### Setting

We then implemented Health Catch-UP! in two urban London primary care practices located in the boroughs of Islington and Brent between September 2021 and March 2022. Sites in these boroughs were selected on two criteria; study interest following participation in the phase one qualitative study and high proportion of migrant (defined as foreign born) residents (Brent: estimated to be 57.0% of population, Islington: estimated to be 42.5%, according to 2021 Census data(29)). Both rank in the top 20% of most deprived local authorities in England, based on the English indices of deprivation 2019.(30)

#### Implementation Support

Training sessions for designated staff working on the study at both sites were completed. Training covered a summary of relevant migrant health screening and vaccination guidelines used in Health Catch-UP!, an introduction and “how to” session for the Health Catch-UP! tool and data collection, and research training that included; good clinical practice, General Data Protection Regulation (GDPR) and ethics. Staff were then supported to download and install the Health Catch-UP! tool onto site computers.

#### Recruitment and Sampling Strategy for Patient Participants

Eligible patients were recruited from the two participating sites. PICOTS criteria are shown in Box 1 below. Eligibility criteria included being aged 18 years or over, a migrant (defined as born overseas), who had moved to the UK at any point, and were able to give informed consent for the study. This broad sampling approach was chosen to test the programme theory’s assumption that Health Catch-UP! would be acceptable to a broad range of migrant groups.

###### Box 1. PICOTS criteria

| **Box 1. PICOTS criteria.** |  |
| --- | --- |
| **Patients** | Adult migrant patients (≥18 years), born outside of United Kingdom. |
| **Intervention** | Clinical Decision Support Tool **Health Catch-UP!** prompting screening and vaccination according to UK guidance |
| **Control** | None |
| **Outcomes** | Number of patients recruited |
|  | Route of recruitment<br>Demographic data (age, sex, country of origin, ethnicity, length time in UK, BMI)<br>Rates of screening offer<br>Uptake of screening offer<br>Number of new conditions diagnosed<br>Rates of under-vaccination for routine vaccine preventable diseases<br>Uptake of routine vaccinations offered<br>Acceptability, feasibility views, barriers and facilitators from practice staff and patients. |
| **Time** | Up to 7 months |
| **Study design** | Mixed-methods process evaluation, two primary care practices London, UK |

Information about the study was translated into the dominant local languages (Arabic, Farsi, Somali and Urdu) and made available to potential participants. At each site, the planned ‘remote’ recruitment approach was initially via a database search for eligible patients who were then contacted using a text message containing a link to a website with further information and the opportunity to express interest in the study. This was unsuccessful in recruiting patients so was superseded by ‘opportunistic’ recruitment in which patients already receiving a face-to-face consultation by a clinician were offered Health Catch-UP! assessment. Patients were given time to read the participant information sheet in their chosen language, telephone interpreters were available on request, and written informed consent was obtained from all participants.

After entry to the study, six demographic questions (ethnicity, age, sex, BMI, country of origin, date of entry to the UK) were coded into Health Catch-UP! within EMIS and integrated into the case records for the patient. For each patient Health Catch-UP! then made suggestions for screening and catch-up vaccination based on the UK guidelines. These were discussed with patients, and the care pathway, as outlined in **Appendix 3** was followed. Where possible, the blood tests for screening and the first doses of a vaccination schedule were planned to be included or booked during the initial appointment.

#### Data Collection, Extraction and Analysis

We sought to collect data relating to the context, mechanisms, and outcomes of Health Catch-UP! implementation to inform formative evaluation and iterative refinement. In line with realist evaluation principles to confirm, refute and refine aspects of our programme theory, we collected qualitative data through interviews with both PHCPs and patients to explore their perspectives on how Health Catch UP! worked in their context. (14) These qualitative data were triangulated with data from the use of the Health Catch-UP! tool in EMIS including quantitative indicators of feasibility and acceptability, outlined below.

Quantitative data collection included:

- Patient demographics: age, sex, BMI, country of origin, ethnicity, date of entry to the UK.
- Recruitment rates by opportunistic and remote routes, including numbers who declined, accepted, and booked an appointment, or accepted but did not attend.
- Rates of screening tests and vaccinations recommended.
- Uptake of screening and vaccination up by patients.
- Number of new conditions identified.

Quantitative data from migrant patient participants enrolled into the study were downloaded from EMIS using a custom-built search into Microsoft Excel. Data were anonymised and securely transferred to the University for analysis in STATA 15. Data cleaning and analyses were done using Microsoft Excel and STATA 15. We used descriptive statistics to describe the demographic characteristics, recruitment, screening and vaccination offer, uptake, and results of participants. We summarised continuous data with mean and standard deviation (SD) and described categorical responses using the frequency and percentage.

Exploratory semi-structured qualitative interviews supported by collaboratively developed topic guides were undertaken with migrants and staff at both sites by SH and LG. Written consent was taken prior to interviews and comprehensive fieldnotes taken (SH and LG) during each interview. These were analysed deductively by hand, according to themes for evaluation of complex interventions recommended by the MRC: acceptability, appropriateness and feasibility.(16) Further qualitative data collection had originally been planned, however this did not go ahead due to the burden of the study upon both sites during the height of the COVID-19 pandemic.

#### Ethics and PPIE

This study received ethics approval from the Health Research Authority and Health and Care Research Wales (IRAS 290630 reference 21/LO/0299), St George’s, University of London Research Ethics Committee (2020.00630) and the Health Research Authority (REC 20/HRA/1674). Migrants with lived experience of the UK immigration and healthcare systems were involved in the design of this study through our National Institute for Health and Care Research (NIHR) funded Patient and Public Involvement and Engagement (PPIE) Project Advisory Board and were compensated for their time and contributions.

## Results

### Phase 1: Iterative Development of Intervention and Initial Programme Theory

#### Initial Programme Theory

The programme theory (**Figure 1)** was developed and iteratively refined collaboratively with stakeholders to provide a visual depiction of our working assumptions regarding the expected inputs, activities, outputs, outcomes and impact of the new pathway, alongside the context, assumptions and unintended consequences (positive and negative).

**Figure 1:**
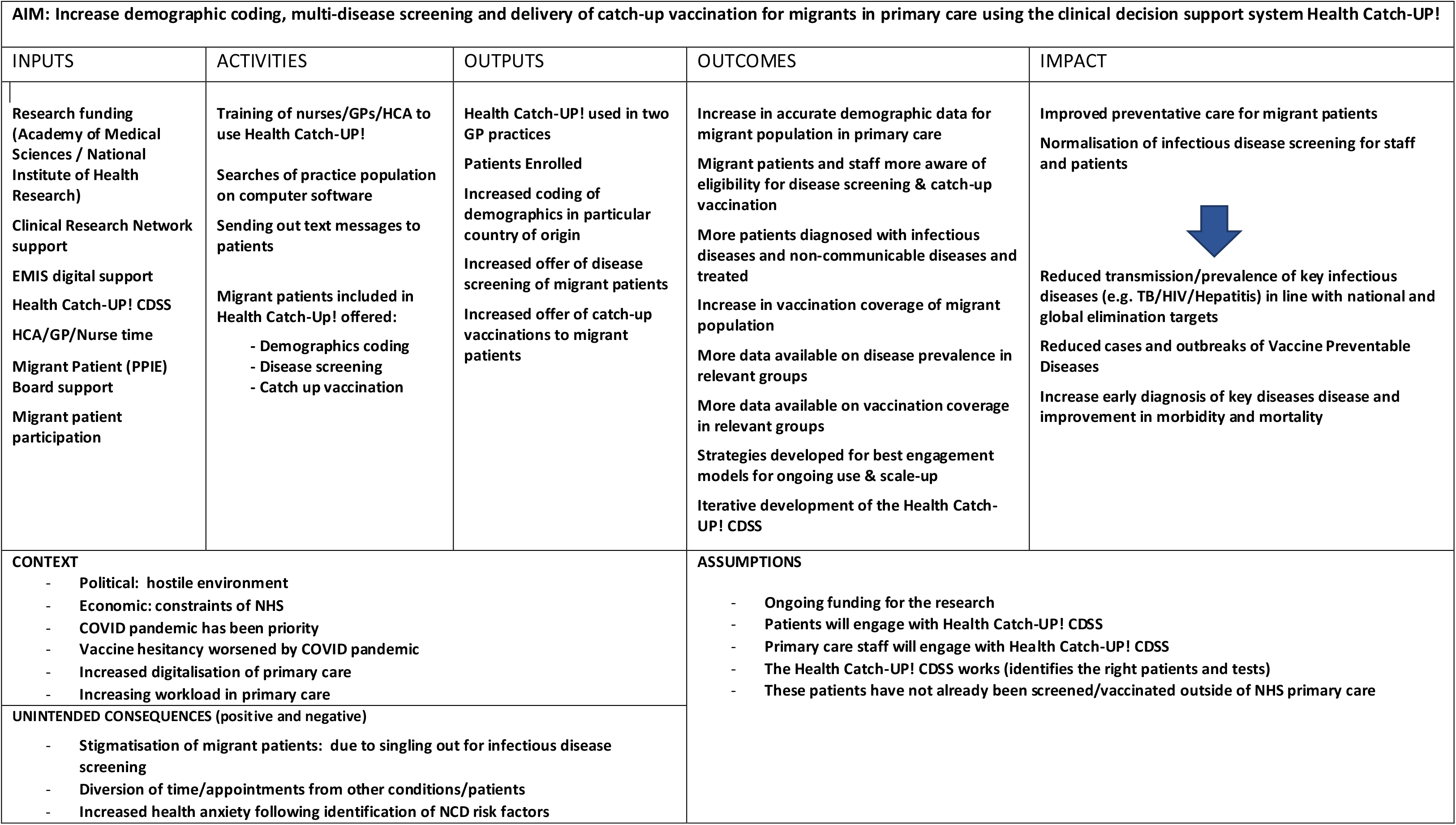
Initial Programme Theory for Health Catch-UP!

#### Key Findings from Phase 1 Qualitative Interviews with Primary Care Practitioners

We interviewed 48 clinicians (25 GPs, 15 practice nurses, 7 health care assistants [allied health professionals who support primary care doctors], 1 pharmacist) and 16 administrative staff (11 Practice-Managers, 5 receptionists). Respondents reported poor implementation of existing screening programmes (such as latent TB) citing overly complex time-consuming pathways without financial and expert support. They felt current infectious disease screening in primary care was not standardised and poorly delivered but could improve with appropriate training and support. Health Catch-UP! was seen as an opportunity to systematically integrate data and support clinical decision-making, and normalisation of primary care-based infectious disease screening for migrants.

Benefits and concerns about Health Catch-UP! were reported at the patient, staff and system level. At the patient level, clinicians felt Health Catch-UP! could provide a ‘one-stop shop’ for preventative healthcare, but reported concerns around the risk of increased stigma, and that patients might be reluctant to share sensitive data (such as country of origin and time in the UK). At a staff level, clinicians felt that Health Catch-UP! would support clinical decision making by providing all the information about the tests the patient was eligible for in one place and therefore reduce workload. However, clinicians recognised that currently these tests are not being generally offered despite patient eligibility, so were concerned about the potential increase in workload and cost of offering and processing additional tests. They reported a lack of knowledge and confidence about how to communicate and manage positive results for infectious diseases. Some staff also reported existing frustration with the number of pop-ups and alerts encountered on EMIS which would be exacerbated by Health Catch-UP! At a system level, perceived benefits included enhancement of health equity and data quality, alongside development of a systematic, standardised approach to screening and catch-up vaccination. Concerns related to incentivisation for potential additional work and whether existing records would provide adequate data to enable identification of migrant patients eligible for screening or vaccination. The full results of this study have been published separately.(9)

These findings led to refining the programme theory and informed our implementation approach in Phase 2 – for example ‘*increased use of appointments’*, was addressed by ensuring Health Catch-UP! could also be delivered opportunistically, which proved critical for recruitment. ‘*Lack of confidence in infectious disease and migrant health*’ was addressed through staff training and the explanation of and signposting to the guidance embedded in the automated features of Health Catch-UP!. The concern around “*pop up fatigue*” was addressed through the CDSS prompts being able to be turned off and used simply as a template.

### Phase 2: Pilot Implementation and Evaluation

#### Implementation

Initial information regarding the requirements of being a research site in this study and research training was provided as planned during the preparation stage. However, subsequently, due to clinical pressures resulting from the pandemic, the decision was made to provide short presentations explaining Health Catch-UP! and how it should be used clinically within existing practice meetings to inform the multi-disciplinary team about the Health Catch-UP! CDSS, rather than providing training at a time that would have taken staff away from their clinical duties. Both sites required support to ensure that they were able to procure all the infectious disease tests required from their core laboratories. However, despite all tests having been initially being set up for procurement, due to the COVID-19 pandemic and difficulties getting results screened at laboratories due to the burden of laboratory workload, the parasitic infections component Health Catch-UP! was turned off.

We recruited 104 participants across two sites, of whom data was available for 99 participants as five participants left the practice before end of the study so data could not be extracted. Most participants (92.92%) were recruited at Site one. The study was open for recruitment between September 2021 and March 2022. Initial recruitment was slow, and the ‘remote route’, in which potential participants were contacted by text message was unsuccessful at recruiting participants. A second wave of recruitment therefore used an opportunistic approach, in which a trusted member of staff introduced the study to potential participants in a routine clinical appointment. At site one, the clinician opportunistically recruiting was the patient’s registered general practitioner, and site two this was a health care assistant. Recruitment is summarised in the flow chart in figure 2.

**Figure 2:**
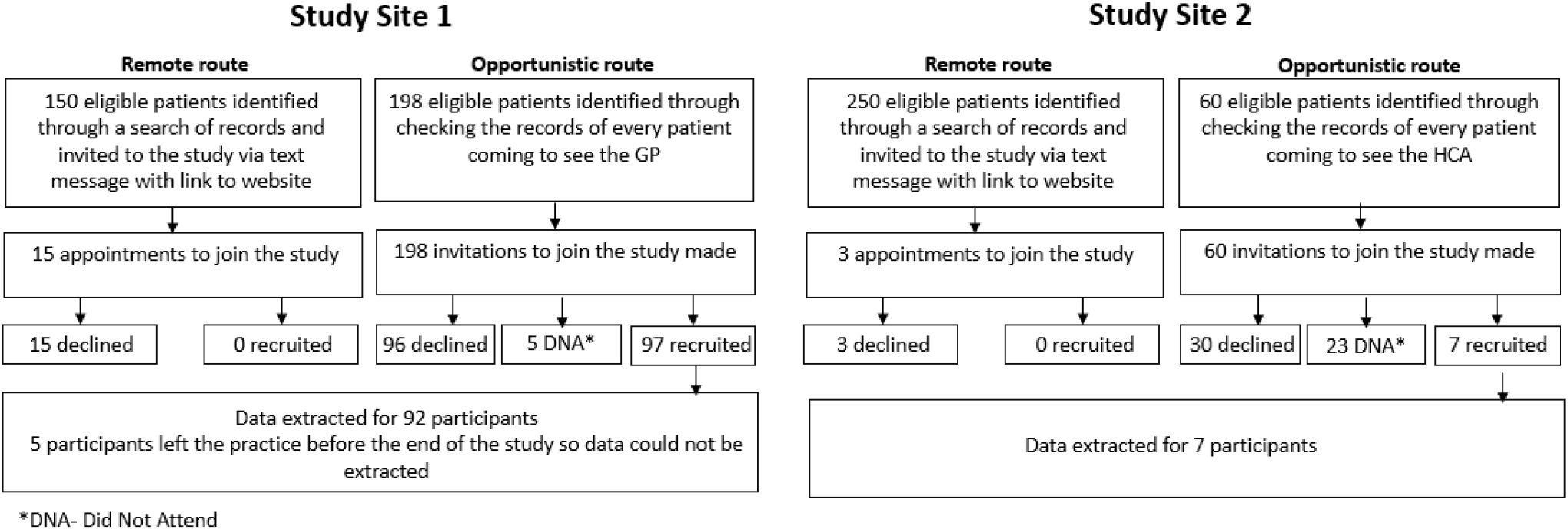
Flow Chart of Patient Recruitment.

#### Outcomes of demographic data collection

Results showed that 100% (n=99) of participants at baseline did not have their country of origin or date of entry to the UK recorded in their primary care records. Health Catch-UP! facilitated completed demographics coding of 96.0% (n=97) of the study population (two participants study data were missing when data were transferred to the research team). Participants were predominantly born in Asia (31.3%, n = 31), followed by Africa (25.2%, n= 25). Further details of country of origin are shown in **Figure 3**. The most common ethnic groups across both sites were Black African/Caribbean (41.41%; n = 41) and Bangladeshi/Indian/Sri Lankan/Pakistani (26.26%; n = 26). Patients at site one were older than at site two with a mean age 60.6 years (SD 14.26) and there was even representation of genders, 48.9% female (n=44). Site one participants had spent longer living in the UK, mean of 33.36 years (SD 19.43). At site two mean age was younger at 39.4 years (SD16.97), participants were predominantly male 85.7% (n=6) and had spent less than 10 years living in the UK (mean years in the UK 8.33, SD 3.22). The study population demographics are summarised in full in **Table 2**.

**Figure 3:**
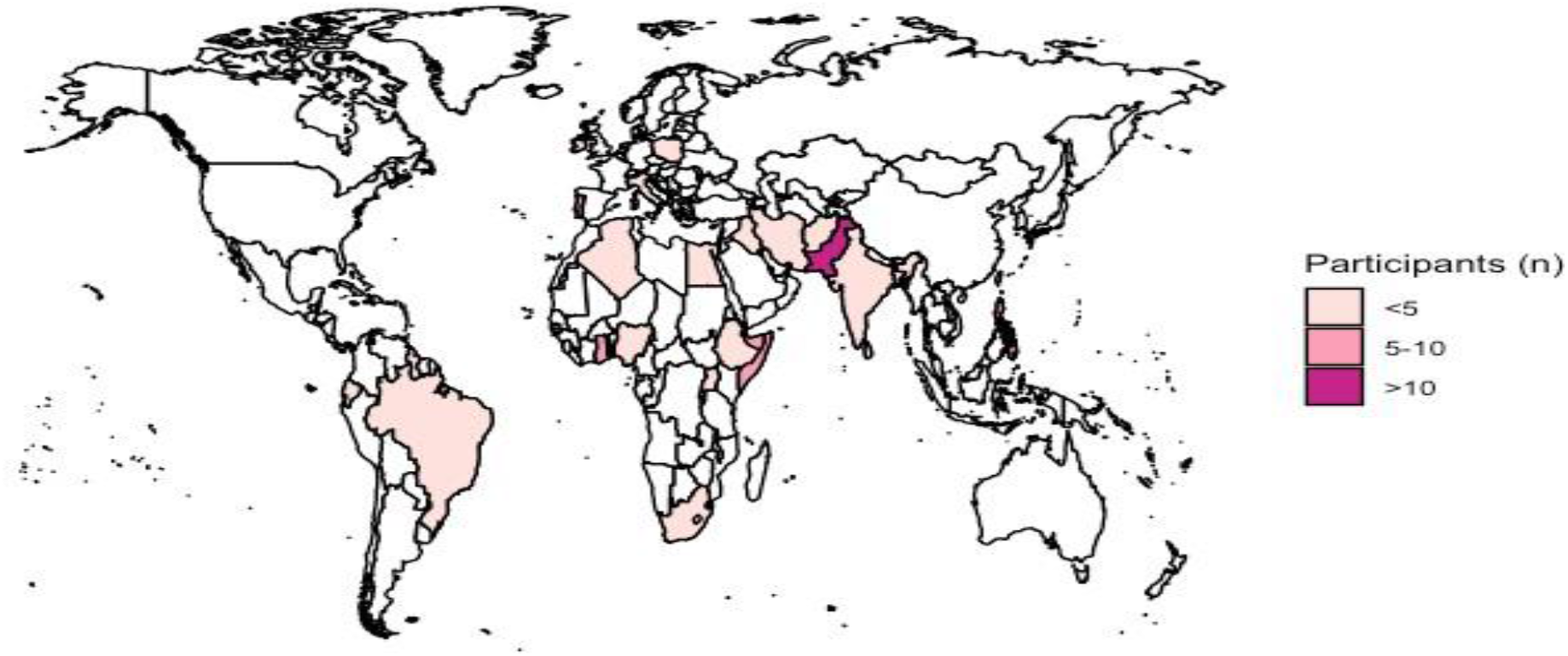
Country of Origin for Recruited Patients.

**Table 2:** Key Features of the Study Population.

|  |  | Site 1 | Site 2 | Total |
| --- | --- | --- | --- | --- |
| Number of patients |  | 92 | 7 | 99 |
| Missing data (n, %) |  | 2 (2.17) | 0 | 2 (2.02) |
| Age (years) mean (SD) |  | 60.60 (14.26) | 39.43 (16.97) | 59.10 (15.37) |
| Sex female, male (% female) |  | 44, 46 (48.89) | 6, 1 (85.71) | 50, 47 (51.54) |
| Ethnicity (n, %) | White UK/Irish | 0 | 0 | 0 |
| Other White |  | 13 (14.13) | 3 (42.86%) | 16 (16.16) |
| Black African/Caribbean |  | 37 (40.22) | 4 (57.14%) | 41 (41.41) |
| Bangladeshi/Indian/Sri Lankan/Pakistani |  | 26 (28.26) | 0 | 26 (28.26) |
| East Asian/Southeast Asian |  | 4 (4.35) | 0 | 4 (4.35) |
| West Asia and North Africa |  | 6 (6.52) | 0 | 6 (6.52) |
| Southern Europe |  | 0 (0.00) | 0 | 0 (0.00) |
| Latin America |  | 2 (2.17) | 0 | 2 (2.17) |
| Mixed Ethnicity |  | 2 (2.17) | 0 | 2 (2.17) |
| Ethnicity recording refused by patient |  | 0 (0.00) | 0 | 0 (0.00) |
| Body Mass Index kg/m<sup>2</sup>, mean (SD) |  | 29.48 (6.29) | 25.31 (4.12) | 29.19 (6.24) |
| Years in the UK mean, SD |  | 33.36 (19.43) | 8.33 (3.22) | 31.59 (19.82) |
| Spent more than 6 months in a high incidence TB country (see Table 1 for definitions) in the last 4 years (n, %) |  | 5 (5.43) | 0 (0) | 5 (5.43) |

#### Outcomes of screening and catch-up vaccination offer

Aggregated data for screening offer, uptake and results across both sites are presented in **Table 3**. The data show that according to UK guidelines, almost two thirds of migrant participants (61.6%, n= 61) were eligible for screening for at least one condition which they had not previously been coded as being offered. Of note 5% (n=5) of the study population were eligible for Latent Tuberculosis (LTBI) screening but had not previously been screened, suggesting that they had been missed by the National LTBI Screening Programme. Of those that were eligible for any screen the majority took up the screening offer (uptake: 86.9%, n = 53) indicating good acceptability of Health Catch-UP!. Viral hepatitis B and C were the most common infectious diseases that participants required screening for with over 40% (n=42) offered hepatitis B screening test and over a third requiring a hepatitis C screen (39.39%; n=39). Of the non-communicable disease screening offered, just under a quarter of patients were eligible for haemoglobinopathy screening (24.24%, n=24), 22% required a cholesterol screening (n=22) and 13.13% a diabetes screen (n=13.13), likely reflecting the age range and raised BMI of the study population, putting them in a higher risk group for these cardiovascular risk factors.

**Table 3:** Outcomes of Screening Process.

|  | **Suggested by**<br>**Health Catch-**<br>**UP!** | **Declined** | **Did not**<br>**attend (DNA)** | **Screened** | **Positive** |
| --- | --- | --- | --- | --- | --- |
| **Infectious disease screening** |  |  |  |  |  |
| Latent TB | 5 (5.05%) | 0 (0.00%) | 1 (20.00%) | 4 (80.00%) | 0 (0.00%) |
| HIV | 11 (11.11%) | 3 (27.27%) | 4 (36.36%) | 4 (36.36%) | 0 (0.00%) |
| Hepatitis B | 42 (40.42%) | 1 (2.38%) | 4 (9.52%) | 37 (88.10%) | 0 (0.00%) |
| Hepatitis C | 39 (39.39%) | 3 (7.69%) | 12 (12.12%) | 24 (30.77%) | 1 (2.56%) |
| **Non-communicable disease screening** |  |  |  |  |  |
| Haemoglobinopathy | 24 (24.24%) | 2 (8.33%) | 7 (29.17%) | 15 (62.50%) | 0 (0.00%) |
| Diabetes | 13 (13.13%) | 0 (0%) | 6 (46.15%) | 7 (53.85%) | 5 (5.05%) |
| At risk of diabetes |  |  |  |  | 4 (30.77%) |
| Suspected diabetes<sup>‡</sup> |  |  |  |  | 1 (7.69%) |
| Cholesterol<sup>¶</sup> | 22 (22.22%) | 0 (0.00%) | 1 (4.55%) | 21 (95.45%) | 6 (27.27%)<sup>¶</sup> |
| **Screening summary** |  |  |  |  |  |
| Recommended at least one screening | 61 (61.61%) |  |  |  |  |
| Attended at least one screening | 53 (86.89%) |  |  |  |  |

| **Catch-up vaccination** |  |
| --- | --- |
| No recorded immunisation | 99 (100%) |
| Offered catch-up vaccination | 99 (100%) |
| Accepted at least one catch-up vaccination | 2 (2.02%) |
| Accepted MMR vaccination | 2 (2.02%) |

<sup>π</sup>At risk of diabetes is defined as HbA1c of (6-6.4%)
<sup>¥</sup>Suspected diabetes is defined as HbA1c of 48mmol/mol (6.5%) or higher
<sup>℥</sup>High cholesterol is defined as total cholesterol of 5 mmol/litre or higher and therefore at risk of cardiovascular disease, needs clinical/lifestyle management

As a result of Health Catch-UP!, 12 new conditions were diagnosed, representing 12.12% of study population and almost a fifth of those eligible for any screening test (19.67%, n = 12, screened total = 61). New diagnoses included hepatitis C (n=1) and eleven non-communicable diseases or risk factors; hypercholesteraemia (n= 6), pre-diabetes (n=4) and diabetes (n=1) again likely reflecting the older age and raised BMI of participants in the study.

The entire study population (n=99) were identified by Health Catch-UP! as being incompletely vaccinated, unvaccinated, or with uncertain vaccination status according to UK immunisation guidelines and required follow up from the practice nurse. (31) This high proportion may reflect genuine under-immunisation or a lack of vaccination data coded into the EMIS system. All participants should then have been offered catch-up vaccination prompted by the Health Catch-UP!, in line with UK guidelines, but uptake was poor with only two participants accepting and receiving MMR vaccination during the study period highlighting that much more needs to be done to support PHCPs with delivering catch-up vaccination to adolescent and adult migrants.

#### Qualitative findings

We interviewed four clinical PHCPs and four patients across both study sites to explore mechanisms of action of Health Catch-UP!, and perception of end-users on the appropriateness, acceptability and feasibility of the tool, and impact of the study’s context. These data are outlined below.

Participant responses to those receiving and using the intervention were positive. PHCPs reported that Health Catch-UP! was generally appropriate and easy to use. Patients reported that being asked in for this check-up felt appropriate for their healthcare and overwhelmingly positive, particularly when offered by a known PHCP. However, further work is needed to understand why the remote route of recruitment via text messaging was so unsuccessful and whether the limited uptake was due to issues with the technology, wording, or external factors such as the ongoing COVID-pandemic and rapid digitalisation of primary care.

In Phase 1, concerns had been raised about patients feeling singled out or discriminated against due to the risk stratification demographic questions required by Health Catch-UP!. In general, PHCPs and patients alike reported that this wasn’t a problem, but that the specific question on length of time in the UK (required for Latent TB infection screening), often elicited strong reactions. This is in line with our previous findings in Phase 1 regarding the difficulties of delivering the National LTBI programme. Concerns around the acceptability of the Health Catch-UP! Process (through collection and coding of demographic data) were largely allayed by effective communication of risk by the PHCP offering the screening.

> *‘However, the “when did you arrive question” was a problem – some were saying vague things, ‘I’ve been here a few years’, others gave an exact date. Some were a bit taken aback – why do you want to know when I arrived here? It’s not routine to ask this question [for Latent TB Infection] at the New patient health check. – HCA, Site 1*

> *‘Because I’ve seen one of the patients was asking ‘Why are you asking me [about my ethnicity]?’ and it was a bit uncomfortable. But the way she explained it, really nice. She was taking her time, sitting with the guy…. I really appreciate it.’ – Patient 4*

Participants felt this was a feasible intervention for primary care to deliver. Both PHCPs and patients commented on its suitability for integration with existing health checks (such as the NHS patient health check and the Over 40s health check) to provide a more comprehensive screen within longer appointments with a preventative health care focus. However, PHCPs felt this would require additional funding, particularly for high-migrant areas. One PHCP felt that migrant groups DNA more than other groups which might affect uptake and recruitment, with cost implications. Another implementation barrier was the logistics of getting tests not routinely done (eg, for parasitic infection and LTBI/IGRA) to the laboratory in time to ensure good sample quality.

> *‘I think GP practices will need to be paid to do this – we already have targets for a new patient health check, so they get paid to do them – I think about 75/85 pounds to the practice – they pay well. But there is an issue in high migrant areas, as their health checks will cost more if you add health catch-up to it.’ – GP, Site 2*

> *“Another barrier is the cut off for lab. We are not big enough for later couriers….but other practices have a phlebotomy service and can get bloods done in the afternoon.’ - HCA, Site 1*

Professional and patient views of Health Catch-UP’s! appropriateness, acceptability and feasibility are expanded upon below in **Table 3**.

**Table 3:** Qualitative Data Pertaining to Appropriateness, Acceptability and Feasibility.

| **Evaluation Domain** | **PHCP comments** | **Patient comments** |
| --- | --- | --- |
| **Appropriateness**<br>Health Catch-UP! is considered appropriate for NHS primary care delivery by professionals and patients, and 'in line' with current work. | *'The tool is straightforward and easy to follow. Because we do NHS checks and diabetes checks we are used to following these types of tools/templates. Putting in ethnicity, bmi, country of origin, they're all asked normally in the new patient health check' – HCA, Site 1* | *'I had a very positive thing. It's a good thing to do. The people who come from abroad, we're born in other countries, so it's a very good thing to check everything for us, blood tests... Also, the injections.... It's a very good experience. You think somebody cares about the people who come from abroad...' – Patient 4* |
| **Acceptability**<br>Health Catch-UP! is acceptable to patients if well-communicated, but the question about date of arrival to the country causes tension. | *'Lots of patients are really keen to get health checks at the moment..... but when we ask questions about when they came into the country, then there's tension. ' – GP, Site 2* | *'I'm not worried about [being offered testing based on my country of origin]. I'm from Pakistan. Some people might generally feel like they're being singled out perhaps, I don't feel I'm being discriminated against if that makes sense...they really took the time to explain that certain ethnicities are more at risk for this reason. And when they explain sometimes the science behind it, you're like, oh, okay, that's why they're doing it. It's not because I'm discriminated against. So, communication is really important there.' – Patient 2* |
| **Feasibility**<br>Health Catch-UP! is considered feasible to be delivered within an existing health check with appropriate resourcing and logistical support. | *'So I think it is a really good idea to do this through the New Patient Health Check. Just going after them and saying – come for your vaccines is not right approach. [It would take] no more than 5 extra minutes for the imms, bloods we are already doing anyway.' – HCA, Site 1* | *'A new patient health check would probably be the best time to get it done and probably would put less stress upon the screening. But in another consultation, 'Oh, by the way, this is what's available for you', just letting them know, it's not bad either. But if I've come in for, say, chest pain or I broke a leg, for example, that's probably the last thing I want to hear.' – Patient 3* |

Identification of any unexpected pathways or consequences of an intervention is a key component of the MRC complex intervention evaluation framework.(17) Clinical staff and the research team at Site one noted unexpected consequences arising from the opportunistic recruitment pathway by the general practitioner. Health Catch-UP! had primarily been designed for the needs of younger migrants who were relatively new arrivals to the country. However, the doctor at Site one noted that by recruiting those who already attending his clinics, he was primarily trialing Health Catch-UP! in an older, more settled cohort of migrant patients. This likely contributed to the significant number of new NCD conditions identified (n = 11) in comparison to the infectious diseases (n=1). There were concerns of further marginalisation of those patients who might have most benefitted, such as refugees and asylum seekers, and labour migrants working longer hours, who may be less likely to access routine primary care during working hours and lack an existing relationship with primary care. These findings highlight the importance of developing flexible and diverse engagement strategies and delivery models to proactively enable vulnerable migrant groups to access Health Catch-UP!.

#### Contextual Changes Over Course of Study

The study was significantly impacted by COVID-19 pandemic and therefore the context was highly atypical for primary care. Health Catch-UP! has a public health, preventative medicine focus, which was significantly deprioritised within the COVID crisis. On a practical level, staff sickness reduced available appointments and patient COVID-related sickness may have impacted attendance at screening appointments. The failure of the remote recruitment route via text messaging may have been directly impacted by the rapid increase in health-related communications received by patients following the rapid digitalisation of primary care. Additionally, several PHCPs believed that the concern and mistrust of the COVID-19 vaccine directly affected vaccine uptake of other vaccines in primary care, and within this study.

> *‘Because the study has overlapped with covid, it’s caused a lot of additional strain on this.’ GP, Site 1 ‘We have just had worse timing in the world for this study, after Covid – people saying with Covid we don’t know what they put into us with Covid and now you are asking for more vaccines in adults – they are adamant they don’t want it.” – HCA, Site 2*

In addition, Health Catch-UP! and its training package was designed prior to the COVID pandemic, and the ensuing rapid digitalisation prompted reflections that to be relevant to the increasingly digital post-pandemic primary care space, Health Catch-UP! needs to be embedded effectively and integrated with new technologies such as translated text messaging and electronic forms that code into the patient’s record directly. This was felt by the research team to be a priority to explore for effective future implementation of Health Catch-UP!

## Discussion

### Key Findings

We successfully engaged two primary care practices in migrant dense areas of London to implement the complex intervention, Health Catch-UP! to support the delivery of evidence-based migrant screening and vaccination recommendations. Implementation of Health Catch-UP! resulted in identification and screening of 99 patients from migrant backgrounds indicating that the Health Catch-UP! tool is feasible, acceptable, and appropriate in this setting. Health Catch-UP! facilitated comprehensive collection and coding of migrant health data, including country of origin and date of entry to the UK in over 97% of participants. This allowed PHCPs to offer multi-disease screening and vaccination “in one go” on an individualised basis grounded in UK primary care-based guidelines.

Across both sites 61.6% (n=61) of participants were eligible for screening for at least one condition which they had not been coded as having been offered. This included 5 participants who were eligible for LTBI screening and who had been missed by the national LTBI screening programme. Once demographic data had been coded, acceptance and uptake of screening was high with over 85% of participants attending for a screen and almost a fifth of those screened (19.67%) subsequently diagnosed with a new condition. Only one of the new conditions diagnosed was an infectious disease, hepatitis C. This is likely reflective of the migrant patients recruited to the study who were older than the migrant groups the research team had had in mind when the tool was initially developed. This unexpected finding prompts the need for implementation models that proactively reach those more vulnerable groups (e.g. asylum seekers, low-skilled labour migrants, those experiencing homelessness) and consideration of including a fuller cardiovascular assessment e.g. adding blood pressure, in line with previous work suggesting migrants and those from black and minority ethnic groups may have worse health outcomes related to non-communicable diseases including diabetes and cardiovascular disease risk factors in primary care. (32–35). 100% of migrant participants were identified as requiring a referral for catch-up vaccination, aligning with previous work showing under-immunisation of migrants in Europe.(36–38).

### Implementation of Health Catch-UP! tool

Implementation of infectious disease and non-communicable disease screening and catch-up vaccination screening in migrant populations is not comprehensively done in UK primary care(2, 8–11, 39) Our study shows that PHCPs support the concept of innovative clinical decision-support systems like Health Catch-UP! to improve effective implementation of screening and vaccination guidance in migrant groups. PHCPs recognised the benefits of adopting this holistic approach to migrant screening, comparing it to similar more established health checks widely implemented in the NHS. Both PHCPs and patients felt Health Catch-UP! was an acceptable, appropriate, and feasible way of implementing national migrant health guidelines on screening and therefore reducing the inequity posed by the current unstandardised status quo. This would in turn, improve early communicable and non-communicable disease detection and protection against vaccine preventable disease in a vulnerable population, in line with global and national government health targets to reduce health inequalities (NHS Long Term Plan) and eliminate key diseases as public health threats (e.g. viral hepatitis).(24, 25, 31, 40) However, our study also found that for Health Catch-UP! to be effective and sustainable it requires logistical support including robust laboratory pathways to ensure ability to access all appropriate screening tests (parasitic diseases and IGRA), further development to improve engagement with offer of catch-up vaccinations, and delivery models ensuring engagement of most at risk patients.

UK primary health care is a diverse and complex landscape requiring flexible interventions, adaptable for use multiple primary care settings. In our evaluation PHCPs were able to change to opportunistic recruitment and ensure delivery by staff from different professional backgrounds. Future work will seek to explore alternative implementation models for the Health Catch-UP! tool in both traditional and alternative primary care settings. Our findings require us to revisit our initial programme theory and consider the development of a Health Catch-UP! implementation package (to be included in *inputs* and *activities* of the programme) to enhance *outputs* (increased screening) and *outcomes* (early disease diagnosis). Intervention package development should involve migrant groups and PHCPs as equal partners to enable effective co-design, building on learning from our evaluation and grounded in lived experiences.(41, 42)

### Contextualisation within Existing Literature

Our findings align with much of the wider literature suggesting that innovative, integrated, cost-effective community and primary care based migrant screening interventions are an essential step to improve migrant health screening and support global and regional elimination targets for key infections. (2, 3, 8, 43–45) However previous screening interventions have largely taken place in secondary care settings with a single disease or speciality focus (e.g., blood-borne viruses, tuberculosis, mental health) and fail to assess risk at an individual patient level. (2, 6, 46–49) Our findings build on the similar IS-MiHealth tool trial in Spain, which suggested that this was feasible and acceptable in primary care settings and improved screening uptake and diagnosis. (50, 51). Similar implementation barriers were uncovered in our Health Catch-UP! study including PHCPs’ knowledge of included infections and vaccinations, and communication of the screening offer in a culturally appropriate way, taking account of language, gender and background (51). On the other hand, preliminary efficacy after implementation of the IS-MiHealth tool were also reported in Spanish primary care, showing a higher screening rate and diagnostic yield for key infections in migrants compared with the routine care. Intervention centres raised their overall monthly diagnostic rate to 5.8 (95% CI 1.2–10.4; *P* = 0.013) additional diagnoses compared with control centres, showing this increase for HIV, hepatitis B, C, tuberculosis, and parasitic infections.(13)

### Strengths and Limitations

A key strength of this study was its innovative approach to a multi-faceted problem, co-developing a CDSS with end-users from the start, based in a digital system that the majority of UK PHCPs use on a daily basis. Our evaluation provided insights into the use of CDSSs for migrant health in primary care in the UK and other host countries, for further refinement before larger scale testing. Conducting this study during the COVID-19 pandemic presented multiple challenges including impacting recruitment to the study, competing primary care priorities, logistical constraints with laboratories and reduction in the qualitative component of the study due to staff time constraints and sickness. The use of text messaging in the context of increased digital health communications, patient reluctance to leave home, and increased vaccine hesitancy may have contributed study recruitment and engagement. It is also likely that the consenting and recruitment procedure, combined with a reluctance to disclose time spent in the UK for many recent migrants (due to concerns about immigration rules and access to healthcare), means the recruited sample is not generalisable to the target population. However, these challenges reflect the realities of offering screening in primary care and provide insights to inform future work on implementation strategies and reasons for engaging and not engaging with Health Catch-UP!.

Future work must build upon existing studies demonstrating cost-effectiveness of screening for each infection,(52–55) to provide at-scale analysis of feasibility, efficacy, and cost-effectiveness, for integration within routine care.

## Conclusion

Our study indicates that an innovative CDSSs like Health Catch-UP! has potential to significantly improve disease detection and delivery of evidence-based screening guidance within primary care for migrant patients. Ensuring that complex interventions such as Health Catch-UP! are effective in real-world settings requires theory informed, co-developed implementation strategies and robust testing and resourcing. Successful adoption of a tool such as Health Catch-UP! In NHS primary care could lead to improved access to care for migrant populations, reduce health disparities, and improve public health though a reduction in the number of people at risk from vaccine-preventable diseases.

## Data Availability

All data produced in the present study are available upon reasonable request to the authors

## Acknowledgements

Migrants with lived experience of the UK immigration and healthcare systems were involved in the design of this study through our National Institute for Health and Care Research (NIHR) funded Patient and Public Involvement and Engagement (PPIE) Project Advisory Board and were compensated for their time and contributions. We thank all the study partners, funders, members of our NIHR Patient and Public Involvement and Engagement Project Advisory Board, and participants for their valuable contributions to this study.

## Authors’ Contributions

SH had the study idea and acquired Academy of Medical Sciences funding. SH and AD designed the study and submitted ethics. JC, FK, LPG, AD, and AC delivered the study, extracted and analysed the data, and wrote a first draft of the paper. SH, JC, SJ, and LPG supported set up and engagement with GP practices and staff throughout the study. All authors contributed to the data interpretation and writing and editing the paper and read and approved the final manuscript. SH is the guarantor of this study.

## Authors’ Twitter handles

Alison Crawshaw: @AlisonCrawshaw

Sally Hargreaves: @sal_hargreaves

Azeem Majeed: @Azeem_Majeed

Lucy Goldsmith: @LucyGoldsmith19

Anna Deal: @Annacadeal

Jessica Carter: @jessicacartergp

Felicity Knights: @faejones

Farah Seedat: @SeedatFarah

Karen Lau: @lau_karen_hk

Sally Hayward: @hayward_sally

Yusuf Ciftci: yusufciftci42 Fatima

Wurie: @WurieFatima

## Availability of data and materials

Data are available on reasonable request from.

## Funding

This study was funded by the Academy of Medical Sciences (SBF005I1) through a Springboard Award to SH. JC is funded by the NIHR and the Wellcome Trust. FK is funded by the NIHR. SH is additionally funded by the NIHR (NIHR300072; NIHR134801), Academy of Medical Sciences (SBF005I1) the La Caixa Foundation (LCF/PR/SP21/52930003), Research England, MRC, and WHO. AM is supported by the NIHR Applied Research Collaboration NW London.

### Declarations

Ethics approval and consent to participate

This study received ethics approval from the Health Research Authority and Health and Care Research Wales (IRAS 290630 reference 21/LO/0299), St George’s, University of London Research Ethics Committee (2020.00630) and the Health Research Authority (REC 20/HRA/1674). All participants provided informed consent to participate.

### Consent for publication

All authors provide consent for publication.

### Competing interests

FW is a member of the Vulnerable Migrants Wellbeing Project Advisory Board, led by the University of Birmingham and Doctors of the World and funded by the Nuffield Foundation. All other authors declare they have no competing interests. The views expressed are those of the author(s) and not necessarily those of the NHS, Department of Health and Social Care, or the NIHR.

## Additional Information

## Appendix 1 MRC Framework on Complex Interventions

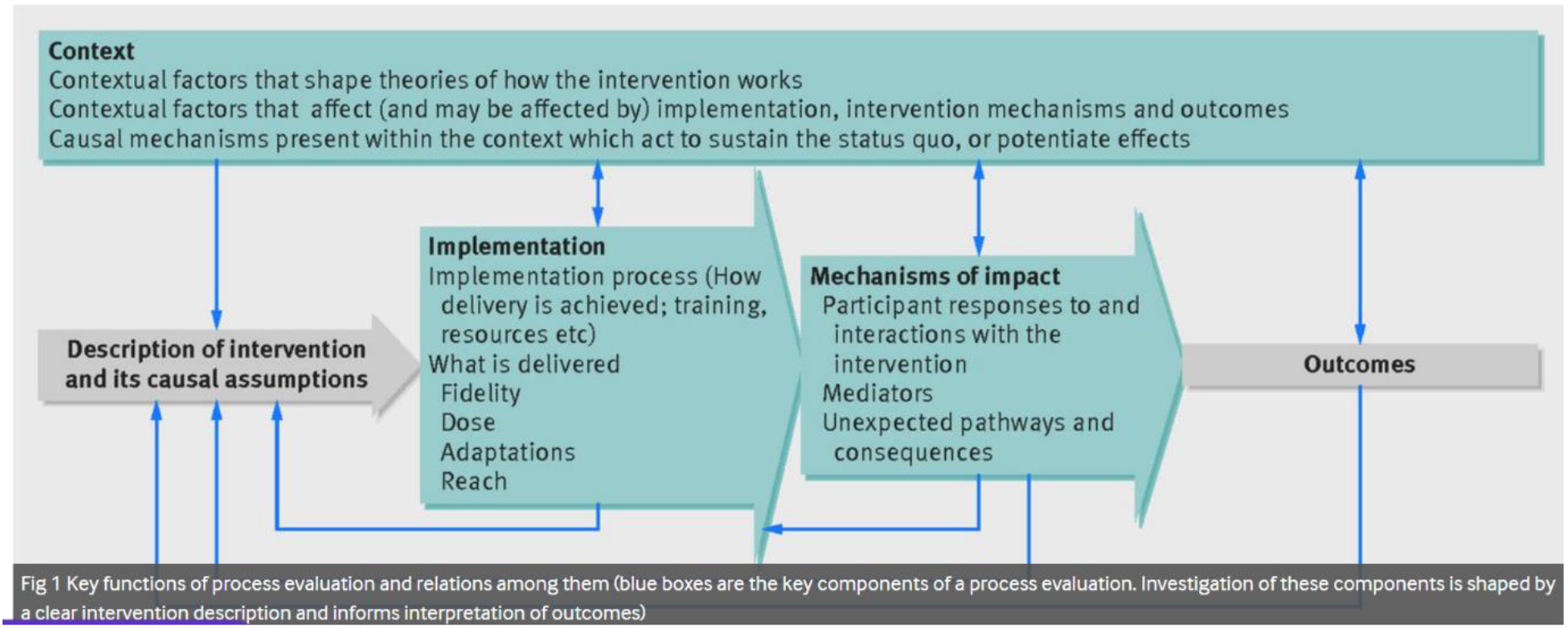

## Appendix 2 Health Catch-UP! Screening and Catch-up Vaccination Prompts

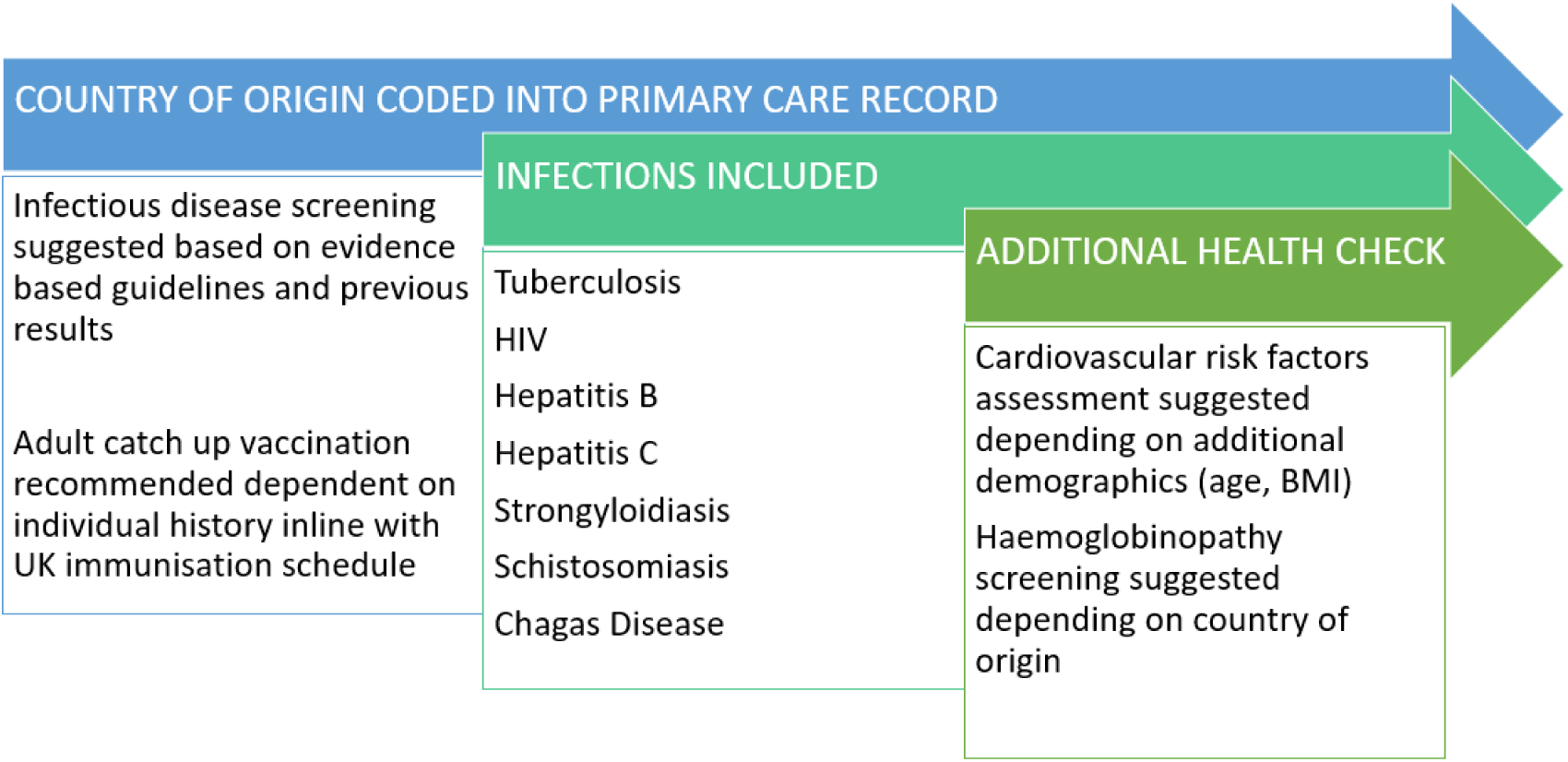

**Health Catch UP! Demonstration link:** https://emishealth.vids.io/videos/a49ad1bb1a18e4c72c/health-catch-up-with-requested-edits-mp4.

## Appendix 3 Health Catch-Up! Care Pathway

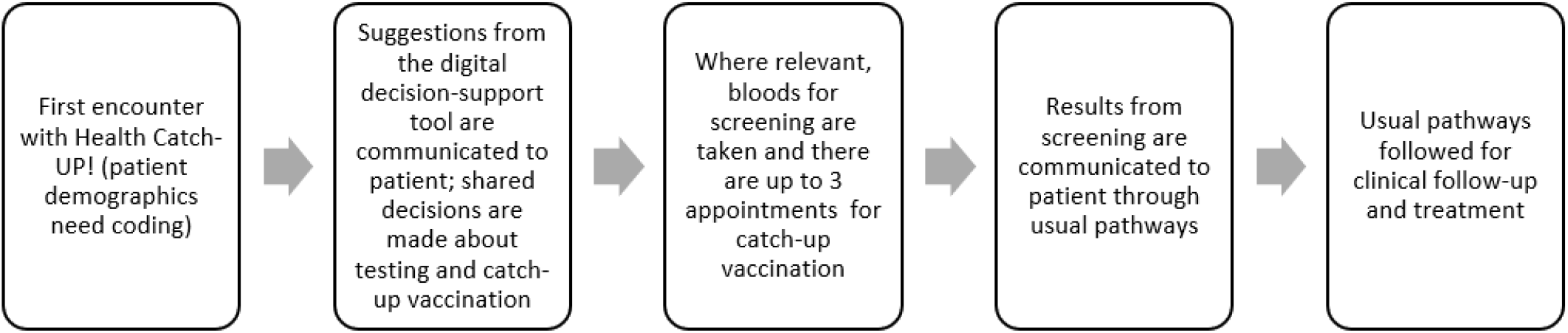

## Notes

### Author Declarations

This study received ethics approval from the Health Research Authority and Health and Care Research Wales (IRAS 290630 reference 21/LO/0299), St George's, University of London Research Ethics Committee (2020.00630) and the Health Research Authority (REC 20/HRA/1674).

